# Use of an extended KDIGO definition to diagnose acute kidney injury in patients with COVID-19: A multinational study of the ISARIC cohort

**DOI:** 10.1101/2022.03.18.22272601

**Authors:** Marina Wainstein, Samual MacDonald, Daniel Fryer, Kyle Young, Steven Webb, Husna Begum, Alistair Nichol, James Lee, Valeria Balan, J. Perren Cobb, Aidan Burrell, Kalynn Kennon, Samantha Strudwick, Sadie Kelly, Malcolm G. Semple, Laura Merson, Srinivas Murthy, Barbara Citarella, Patrick Rossignol, Rolando Claure-Del Granado, Sally Shrapnel, The ISARIC Characterization Group

## Abstract

**Background:** Acute kidney injury (AKI) is one of the most common and significant problems in patients with COVID-19. However, little is known about the incidence and impact of AKI occurring in the community or early in the hospital admission. The traditional KDIGO definition can fail to identify patients for whom hospitalization coincides with recovery of AKI as manifested by a decrease in serum creatinine (sCr). We hypothesized that an extended KDIGO definition, adapted from the International Society of Nephrology 0by25 studies, would identify more cases of AKI in patients with COVID-19 and that these may correspond to community-acquired AKI with similarly poor outcomes as previously reported in this population.

**Methods and Findings:** All individuals in the ISARIC cohort admitted to hospital with SARS-CoV-2 infection from February 15^th^, 2020, to February 1^st^, 2021, were included in the study. Data was collected and analysed for the duration of a patient’s admission. Incidence, staging and timing of AKI were evaluated using a traditional and extended KDIGO (eKDIGO) definition which incorporated a commensurate decrease in serum creatinine. Patients within eKDIGO diagnosed with AKI by a decrease in sCr were labelled as deKDIGO. Clinical characteristic and outcomes – intensive care unit (ICU) admission, invasive mechanical ventilation and in-hospital death - were compared for all three groups of patients. The relationship between eKDIGO AKI and in-hospital death was assessed using survival curves and logistic regression, adjusting for disease severity and AKI susceptibility. 75,670 patients from 54 countries were included in the final analysis cohort. Median length of admission was 12 days (IQR 7, 20). There were twice as many patients with AKI identified by eKDIGO than KDIGO (31.7 vs 16.8%). Those in the eKDIGO group had a greater proportion of stage 1 AKI (58% vs 36% in KDIGO patients). Peak AKI occurred early in the admission more frequently among eKDIGO than KDIGO patients. Compared to those without AKI, patients in the eKDIGO group had worse renal function on admission, more in-hospital complications, higher rates of ICU admission (54% vs 23%) invasive ventilation (45% vs 15%) and increased mortality (38% vs 19%). Patients in the eKDIGO group had a higher risk of in-hospital death than those without AKI (adjusted OR: 1.78, 95% confidence interval: 1.71-1.8, p-value < 0.001). Mortality and rate of ICU admission were lower among deKDIGO than KDIGO patients (25% vs 50% death and 35% vs 70% ICU admission) but significantly higher when compared to patients with no AKI (25% vs 19% death and 35% vs 23% ICU admission) (all p values < 5×10^−5^). Limitations include ad hoc sCr sampling, exclusion of patients with less than two sCr measurements, and limited availability of sCr measurements prior to initiation of acute dialysis.

**Conclusions:** The use of an extended KDIGO definition to diagnose AKI in this population resulted in a significantly higher incidence rate compared to traditional KDIGO criteria. These additional cases of AKI appear to be occurring in the community or early in the hospital admission and are associated with worse outcomes than those without AKI.

**Author Summary:** *Why was this study done?:*

- Previous studies have shown that acute kidney injury (AKI) is a common problem among hospitalized patients with COVID-19.
- The current biochemical criteria used to diagnose AKI may be insufficient to capture AKI that develops in the community and is recovering by the time a patient presents to hospital.
- The use of an extended definition, that can identify AKI both during its development and recovery phase, may allow us to identify more patients with AKI. These patients may benefit from early management strategies to improve long term outcomes.

*What did the researchers do and find?:*

- In this study, we examined AKI incidence, severity and outcomes among a large international cohort of patients with COVID-19 using both a traditional and extended definition of AKI.
- We found that using the extended definition identified almost twice as many cases of AKI than the traditional definition (31.7 vs 16.8%).
- These additional cases of AKI were generally less severe and occurred earlier in the hospital admission. Nevertheless, they were associated with worse outcomes, including ICU admission and in-hospital death (adjusted odds ratio: 1.78, 95% confidence interval: 1.71-1.8, p-value < 0.001) than those with no AKI.

*What do these findings mean?:*

- The current definition of AKI fails to identify a large group of patients with AKI that appears to develop in the community or early in the hospital admission.
- Given the finding that these cases of AKI are associated with worse admission outcomes than those without AKI, identifying and managing them in a timely manner is enormously important.

## Introduction

Acute kidney injury (AKI) has been identified as one of most common and significant problems in hospitalized patients with COVID 19 [1, 2]. Observational studies have consistently shown that patients who develop AKI are more likely to be admitted to an intensive care unit (ICU), require invasive mechanical ventilation, have longer lengths-of-stay (LOS) and increased mortality [1-3]. Autopsy studies point to several potential pathophysiological pathways for AKI including acute tubular injury from hemodynamic shifts, local inflammatory and microvascular thrombotic changes from immune dysregulation as well as direct viral invasion through the angiotensin converting receptor enzyme-2 (ACE-2) receptor [4].

Until now, most studies looking at AKI in COVID-19 have used the traditional Kidney Disease Improving Global Outcomes (KDIGO) definition which relies on the rise in serum creatinine, either by 0.3 mg/dl in 48 hours or by 50% from baseline over a seven-day period [5]. While this definition is likely to adequately capture AKI that develops during a hospital stay, it may fail to identify cases that have developed in the community and are potentially recovering by the time a patient presents to the hospital, thereby underestimating the true incidence of AKI. To address this potential limitation of the KDIGO definition, the International Society of Nephrology (ISN) 0by25 studies added a commensurate fall in serum creatinine to their definition of AKI [6, 7]. Using this modified criterion in the feasibility study, it was found that approximately 40% of the community-acquired AKI (CA-AKI) could be identified by a fall in the level of serum creatinine early in the admission making it a more comprehensive and inclusive definition [7].

The integration of this additional criteria to identify kidney injury has also been highlighted as one of the research priorities in the recent KDIGO report on controversies in AKI [8]. While other papers have indicated the need to revise various aspects of the KDIGO criteria, aside from the 0by25 studies, a decrease in serum creatinine as a marker of AKI has only been explored in infants and neonates, prompting a need for further research in this area [9, 10].

Given the global impact of SARS-CoV-2 infection across all income and resource settings, combined with the potentially significant burden of AKI occurring in the community, we hypothesized that an extended KDIGO definition, adapted from the ISN 0by25 studies, would identify more cases of AKI in patients with COVID-19. We also hypothesized that the additional cases identified using this extended criterion may correspond to CA-AKI and be associated with similarly poor outcomes as those shown in previous studies of AKI in COVID-19 [1-3]

## Methods

### Study Design

The International Severe Acute Respiratory and Emerging Infection Consortium (ISARIC) - World Health Organization (WHO) Clinical Characterization Protocol for Severe Emerging Infections provided a framework for prospective, observational data collection on hospitalized patients. The protocol, case report forms and study information are available online (https://isaric.org/research/covid-19-clinical-research-resources). This protocol builds on the ISARIC/WHO COVID-19 Clinical Characterisation Protocol (CCP) and associated data collection forms already in operation, the Core and Rapid case report forms (CRFs) [11], of which only the core CRF was used in this study. These CRFs were developed to standardize clinical data collection on patients admitted with suspected or confirmed COVID-19. They are used to collect data on demographics, pre-existing comorbidities and risk factors, signs and symptoms experienced during the acute phase, and care and treatments received during hospitalization. Collection of serum creatinine measurements across all sites was not time-standardized and the frequency of collection was left to the discretion of each site.

This observational study required no change to clinical management and permitted patient enrollment in other research projects. Protocol and consent forms are available at https://isaric.net/ccp/. While written consent was obtained in most cases, for some sites the local regulators and ethics committees approved oral consent, or waiver of consent, in the context of the pandemic.

The ISARIC-WHO Clinical Characterisation Protocol was approved by the World Health Organization Ethics Review Committee (RPC571 and RPC572). Ethical approval was obtained for each participating country and site according to local requirements (S1 statement). A prospective analysis plan was used to guide the design of this study [12]. Only the first of the three aims proposed was addressed in this study and the addition of an extended AKI definition was adapted from the 0by25 studies in order to better capture AKI occurring in the community or early in the hospital admission [6].

### Study population

#### Inclusion & exclusion criteria

All individuals in the ISARIC database with clinically diagnosed or laboratory-confirmed SARS-CoV-2 infection admitted to hospital from February 15^th^, 2020, to February 1^st^ 2021 (criteria for clinical diagnosis in S1 Table) were included in the study cohort. Patients younger than 18 years of age and those on maintenance kidney replacement therapy (dialysis or transplantation) were excluded. Patients with fewer than two serum creatinine (sCr) measurements during the admission and those with incomplete or unreliable laboratory data were also excluded (Fig 1).

**Fig 1.**
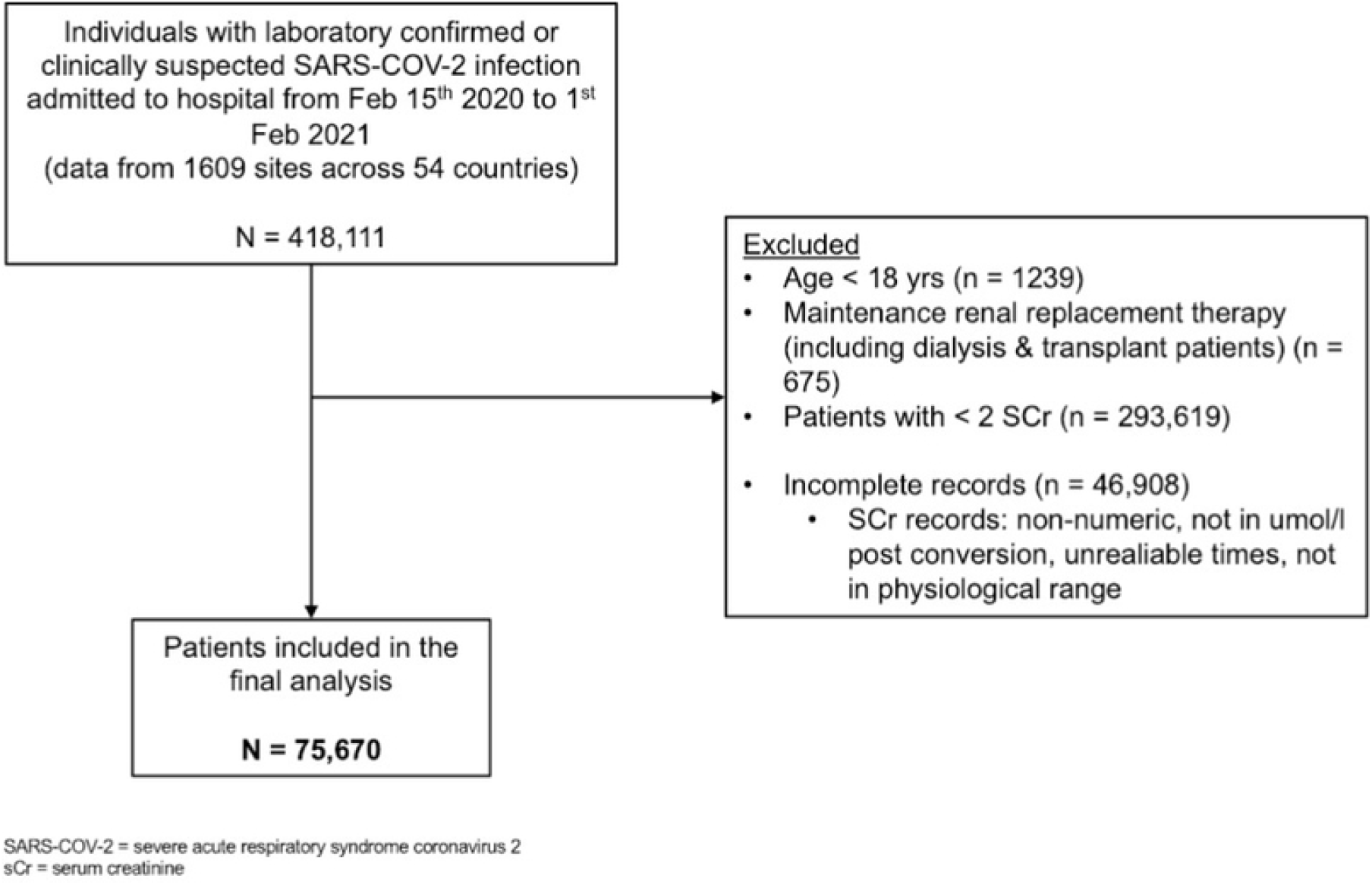
Flowchart of the study

### AKI definition and diagnosis

AKI was identified biochemically using sCr and incidence rates calculated accordingly. Patients’ sCr levels throughout the admission were used to classify them as (i) not having AKI, (ii) having AKI according to the traditional KDIGO definition, or (iii) AKI according to the extended KDIGO (eKDIGO) criteria. For the purpose of this analysis, those patients in the eKDIGO group with AKI diagnosed by a fall in sCr have been labelled as deKDIGO (Fig 2). The KDIGO definition of AKI requires a patient to have an increase in sCr by 0.3 mg/dl within 48 hours or an increase to more than 1.5 times the baseline sCr within 7 days [5]. The eKDIGO definition of AKI was adapted from the ISN’s 0by25 trial and includes a fall in sCr by 0.3 mg/dl within 48 hours or a fall to more than 1.5 times the baseline sCr within 7 days [7]. Acute kidney injury was then graded according to the corresponding staging criteria for each definition (Table 1). A moving window of 48 hours and 7 days was applied during the entire length of a patient’s admission to find the first instance of AKI as well as the highest stage reached. In the case of AKI diagnosis using an increment in sCr, the minimum sCr within that window was deemed as the baseline, while for a diagnosis using the decrement, the maximum sCr within that window became the baseline. Information on the timing of acute RRT was not always available so it was not possible to determine whether patients receiving RRT fell into the KDIGO or deKDIGO portions of the eKDIGO group. Given the low likelihood that a patient with a falling creatinine would be given acute RRT, all RRT patients were categorized as being stage 3 AKI within the KDIGO group. Urine volume criteria was not used for either definition as it was not routinely collected in the CRF.

**Table 1.**
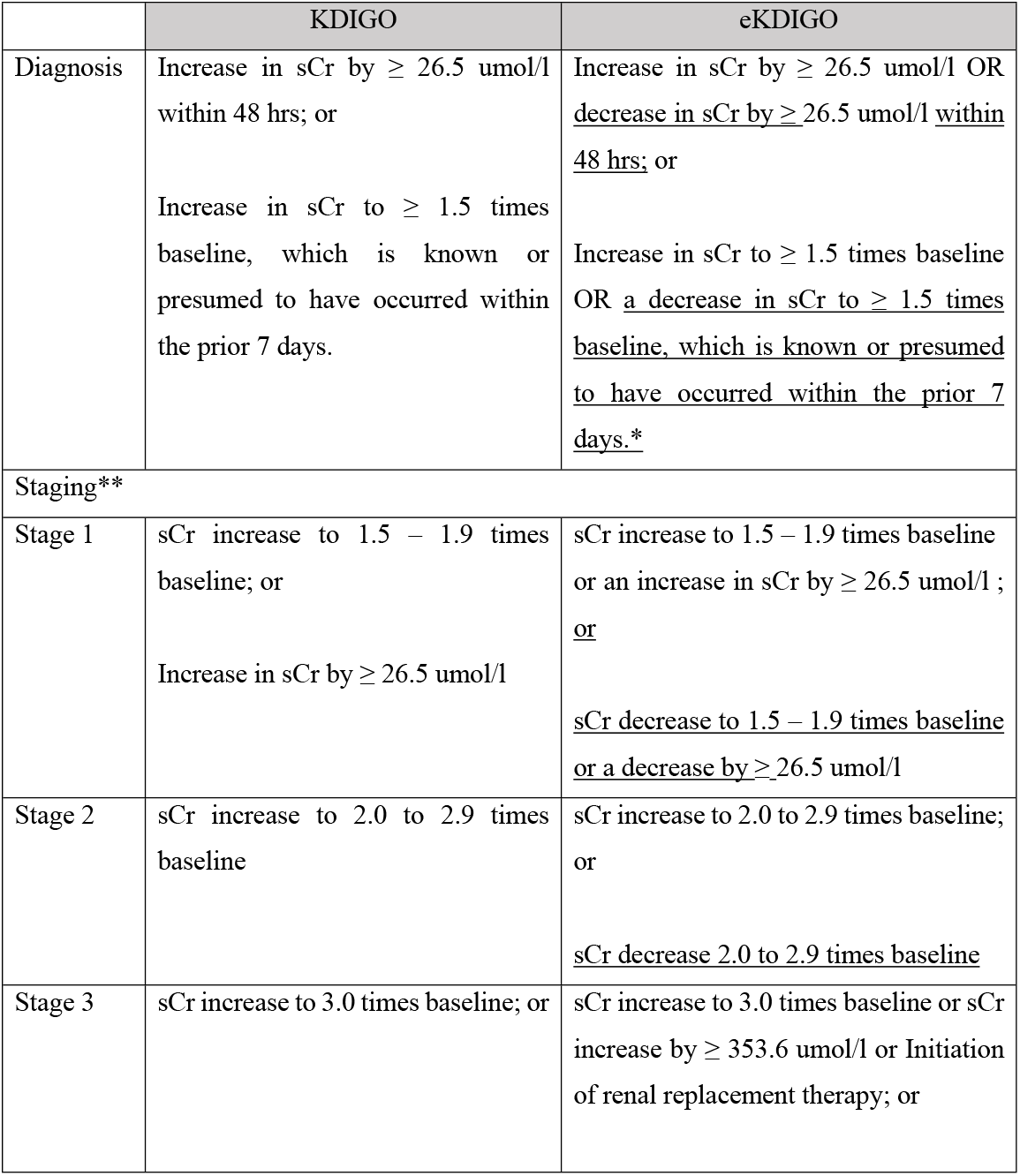
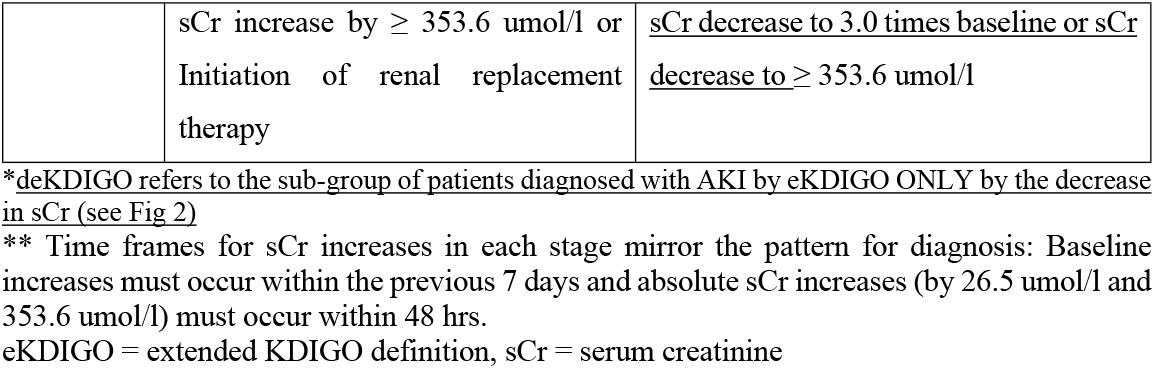
Acute Kidney Injury Definitions.

|  | KDIGO | eKDIGO |
| --- | --- | --- |
| Diagnosis | Increase in sCr by $\geq 26.5$ $\mu\text{mol/l}$ within 48 hrs; or<br>Increase in sCr to $\geq 1.5$ times baseline, which is known or presumed to have occurred within the prior 7 days. | Increase in sCr by $\geq 26.5$ $\mu\text{mol/l}$ OR <u>decrease in sCr by $\geq 26.5$ $\mu\text{mol/l}$ within 48 hrs;</u> or<br>Increase in sCr to $\geq 1.5$ times baseline OR <u>a decrease in sCr to $\geq 1.5$ times baseline, which is known or presumed to have occurred within the prior 7 days.*</u> |
| Staging** |  |  |
| Stage 1 | sCr increase to 1.5 – 1.9 times baseline; or<br>Increase in sCr by $\geq 26.5$ $\mu\text{mol/l}$ | sCr increase to 1.5 – 1.9 times baseline or an increase in sCr by $\geq 26.5$ $\mu\text{mol/l}$ ;<br><u>or</u><br><u>sCr decrease to 1.5 – 1.9 times baseline</u><br><u>or a decrease by $\geq 26.5$ $\mu\text{mol/l}$</u> |
| Stage 2 | sCr increase to 2.0 to 2.9 times baseline | sCr increase to 2.0 to 2.9 times baseline; or<br><u>sCr decrease 2.0 to 2.9 times baseline</u> |
| Stage 3 | sCr increase to 3.0 times baseline; or | sCr increase to 3.0 times baseline or sCr increase by $\geq 353.6$ $\mu\text{mol/l}$ or Initiation of renal replacement therapy; or |
| | sCr increase by $\geq 353.6$ $\mu\text{mol/l}$ or<br>Initiation of renal replacement<br>therapy | <u>sCr decrease to 3.0 times baseline or sCr</u><br><u>decrease to $\geq 353.6$ $\mu\text{mol/l}$</u> |

\*deKDIGO refers to the sub-group of patients diagnosed with AKI by eKDIGO ONLY by the decrease in sCr (see Fig 2)
\*\* Time frames for sCr increases in each stage mirror the pattern for diagnosis: Baseline increases must occur within the previous 7 days and absolute sCr increases (by $26.5 \mu\text{mol/l}$ and $353.6 \mu\text{mol/l}$ ) must occur within 48 hrs.
eKDIGO = extended KDIGO definition, sCr = serum creatinine

**Fig 2.**
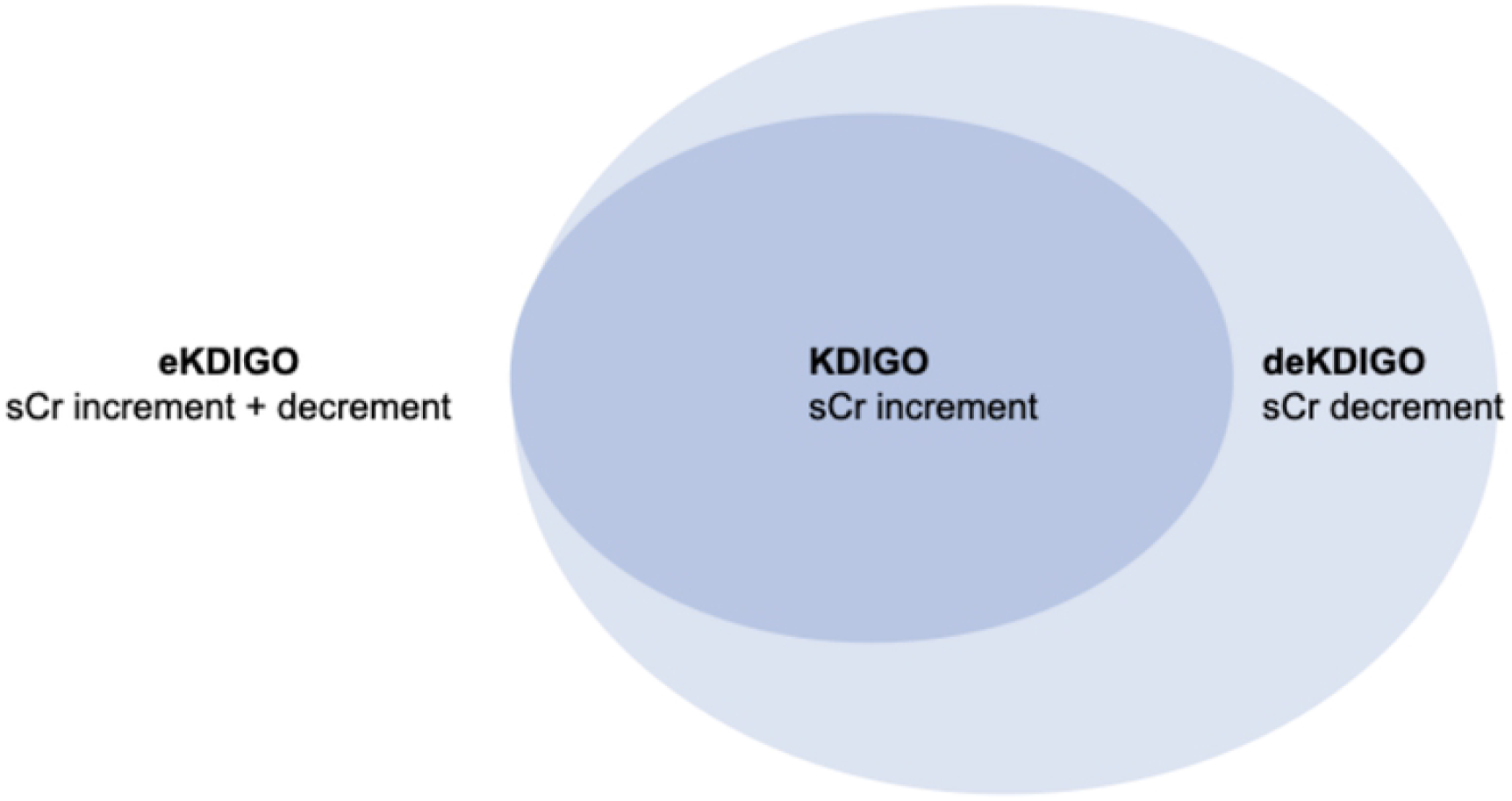
Visual representation of the relationships between serum creatinine trajectories within the AKI groups (KDIGO, eKDIGO, deKDIGO).

### Data collection and time to peak AKI

This study analysed data up until February 15^th^ on patients for whom data collection commenced on or before February 1^st^, 2021. Data was collected and analysed for the duration of a patient’s admission. A 14-day rule was applied to focus analysis on individuals who were more likely to have a recorded outcome. By excluding patients enrolled during the last 14 days, we aimed to reduce the number of incomplete data records, thus improving the generalizability of the results and the accuracy of the outcomes.

For both groups (KDIGO and eKDIGO) time to peak AKI from hospital admission and the respective counts for each day were compared by visual inspection of histograms using the first day that a peak stage was reached. From the pre-specified data collected in the CRF information was obtained on demographics and country income level divided according to the World Bank classification (https://data.worldbank.org/country) into high income (HIC), upper middle income (UMIC) and low and low middle-income countries (LLMIC) merged into a single category. Information was obtained on patients’ comorbidities and pre-admission medications as well as signs and symptoms, observations and laboratory results on admission. Information collected during the admission included acute treatments, complications and outcomes. Outcomes included an admission to the ICU, use of invasive mechanical ventilation, and either discharge, transfer to another hospital, in-hospital death or remaining in hospital. Definitions of all collected variables are provided in S2 Table. A comparison of these variables was performed among patients with eKDIGO AKI and no AKI; deKDIGO and KDIGO AKI; deKDIGO AKI and no AKI.

Patients were classed as lost to follow up if either a) they were transferred to another facility, or b) they had an unknown outcome and the last date upon which any data was recorded for them was 45 days or before the date of data extraction. Patients with unknown outcome where the last recorded data was less than 45 days old are categorized as receiving ongoing care. Data on readmissions could not be obtained for patients in many countries.

### Statistical analysis

For continuous variables, characteristics were reported as medians and interquartile ranges (IQR). For categorical variables, counts and percentages were reported. All statistical tests were carried out as pairwise independent samples comparisons. Due to the number of statistical tests conducted, a conservative Bonferroni adjusted significance level of α_b_ 5 × 10-5 was used to limit the study wide probability of a type I error [13]. For continuous variables, the Mann-Whitney U test was used. For categorical variables, Pearson’s chi-squared test was performed. Missing data was reported as a percentage of the relevant cohort for each variable in Table 2, 3 and 4 and further information on its distribution is presented in S3 Table.

**Table 2.** Characteristics of patients with no AKI and AKI diagnosed by eKDIGO definition.

|  |  | eKDIGO AKI | No AKI | Missingness (%) | P-value |
| --- | --- | --- | --- | --- | --- |
| **Total Count** |  |  |  |  |  |
|  |  | 23892 | 51772 |  |  |
| **Demographics** |  |  |  |  |  |
|  | Age, yr, median (IQR) | 68 (57.5, 78.5) | 67 (53.5, 80.5) | 0 | p < 5x10<sup>-5</sup> |
|  | Female (%) | 8375 (35) | 22031 (43) | 0 | p < 5x10<sup>-5</sup> |
| **Country Income Level, n (%)** |  |  |  |  |  |
|  | High Income | 20513 (86) | 45686 (88) | 0 | p < 5x10<sup>-5</sup> |
|  | Upper Middle Income | 1129 (5) | 4176 (8) | 0 | p < 5x10<sup>-5</sup> |
|  | Low & low middle income | 2161 (9) | 1876 (4) | 0 | p < 5x10<sup>-5</sup> |

| **AKI Grades & RRT, n (%)** |  |  |  |  |  |
| --- | --- | --- | --- | --- | --- |
|  | AKI Stage 1 | 13746 (58) | - | 0 |  |
|  | AKI Stage 2 | 3682 (15) | - | 0 |  |
|  | AKI Stage 3* | 6464 (27) | - | 0 |  |
|  | RRT | 4252 (19) | - | 9 |  |
| **Comorbidities\*\*, n (%)** |  |  |  |  |  |
| | Chronic Kidney Disease | 4059 (18) | 5433 (11) | 7 | $p < 5 \times 10^{-5}$ |
|  | Chronic Cardiac Disease | 6225 (27) | 12772 (25) | 2 |  |
|  | Chronic Pulmonary Disease | 2975 (13) | 6853 (14) | 7 |  |
| | Hypertension | 8447 (50) | 16429 (43) | 28 | $p < 5 \times 10^{-5}$ |
|  | Dementia | 1835 (9) | 4315 (9) | 12 |  |
| | Diabetes -type 2 | 7996 (36) | 14051 (29) | 6 | $p < 5 \times 10^{-5}$ |
|  | Liver disease | 842 (4) | 1653 (3) | 4 |  |
|  | Malnutrition | 501 (2) | 963 (2) | 12 |  |
| | Obesity | 3792 (19) | 6404 (15) | 18 | $p < 5 \times 10^{-5}$ |
| **Medications on Admission, n (%)** |  |  |  |  |  |
|  | NSAIDS | 1382 (9) | 2610 (8) | 35 |  |
| | ACEi | 2795 (17) | 5177 (15) | 33 | $p < 5 \times 10^{-5}$ |
| | ARBs | 1942 (12) | 3101 (9) | 33 | $p < 5 \times 10^{-5}$ |
| **Signs & Symptoms on Admission, n (%)** |  |  |  |  |  |
| | Altered Consciousness / Confusion | 4295 (23) | 8267 (20) | 19 | $p < 5 \times 10^{-5}$ |
|  | Diarrhea | 3740 (20) | 7983 (19) | 19 |  |
|  | Fever | 13333 (64) | 30320 (66) | 12 |  |
|  | Vomiting /nausea | 3387 (18) | 8099 (19) | 19 |  |
| | Muscle aches/joint pain | 3643 (21) | 8853 (23) | 25 | $p < 5 \times 10^{-5}$ |
| | Headache | 1779 (10) | 5367 (14) | 26 | $p < 5 \times 10^{-5}$ |
|  | Sore Throat | 1564 (9) | 3579 (9) | 27 |  |
|  | Cough | 12961 (63) | 29549 (65) | 12 |  |
| | Shortness of breath | 14824 (72) | 30334 (66) | 12 | $p < 5 \times 10^{-5}$ |
|  | Runny nose | 634 (4) | 1547 (4) | 28 |  |
| **Observations on Admission, median (IQR)** |  |  |  |  |  |
|  | Temperature, C | 37.2 (36.2, 38.2) | 37.3 (36.8, 37.8) | 3 |  |
| | Systolic BP, mmHg | 127 (110.5, 143.5) | 130 (115.0, 145.0) | 6 | $p < 5 \times 10^{-5}$ |
| | Diastolic BP, mmHg | 71 (61.5, 80.5) | 75 (66.0, 84.0) | 7 | $p < 5 \times 10^{-5}$ |
| | Heart rate, BPM | 93 (79.5, 106.5) | 90 (77.5, 102.5) | 7 | $p < 5 \times 10^{-5}$ |
| | Respiratory rate, per min | 23 (18.5, 27.5) | 21 (17.0, 25.0) | 13 | $p < 5 \times 10^{-5}$ |
| | Oxygen saturation, % | 94 (90.5, 97.5) | 95 (92.5, 97.5) | 7 | $p < 5 \times 10^{-5}$ |
| **Laboratory Results on Admission, median (IQR)** |  |  |  |  |  |
| | WBC ($\times 10^9/L$) | 8.2 (5.2, 11.2) | 7 (4.5, 9.5) | 15 | $p < 5 \times 10^{-5}$ |
| | BUN (mmol/L) | 10.9 (4.9, 16.9) | 6.4 (3.9, 8.9) | 22 | $p < 5 \times 10^{-5}$ |
| | Potassium (mmol/L) | 4.2 (3.8, 4.6) | 4.1 (3.7, 4.4) | 15 | $p < 5 \times 10^{-5}$ |
| | CRP (mg/L) | 95.3 (19.8, 170.8) | 69 (12.5, 125.5) | 22 | $p < 5 \times 10^{-5}$ |
| | sCr ($\mu\text{mol/l}$) | 110 (67.5, 152.5) | 80 (63.0, 97.0) | 11 | $p < 5 \times 10^{-5}$ |
| | eGFR ($\text{ml/min/1.73m}^2$) | 54.4 (29.4, 79.4) | 80.3 (61.8, 98.8) | 12 | $p < 5 \times 10^{-5}$ |
| **Admission Treatment, n (%)** |  |  |  |  |  |
| | Antiviral and COVID-19 targeting agents | 5349 (26) | 9145 (21) | 16 | $p < 5 \times 10^{-5}$ |
| | Antibiotic agents | 20905 (93) | 40430 (86) | 8 | $p < 5 \times 10^{-5}$ |
| | Antifungal agents | 2459 (11) | 2564 (6) | 11 | $p < 5 \times 10^{-5}$ |
| | Corticosteroids | 8553 (38) | 11905 (25) | 9 | $p < 5 \times 10^{-5}$ |
| **Complications\*\*, n (%)** |  |  |  |  |  |
| | Bacterial pneumonia | 3827 (19) | 5761 (13) | 16 | $p < 5 \times 10^{-5}$ |
| | Cardiac arrest | 1482 (7) | 988 (2) | 10 | $p < 5 \times 10^{-5}$ |
| | Coagulation disorder | 1414 (7) | 1321 (3) | 15 | $p < 5 \times 10^{-5}$ |
| | Rhabdomyolysis | 292 (1) | 177 (0.4) | 15 | $p < 5 \times 10^{-5}$ |
| **Outcomes, n (%)** |  |  |  |  |  |
| | ICU admission | 12579 (54) | 11652 (23) | 2 | $p < 5 \times 10^{-5}$ |
| | Invasive mechanical ventilation | 10264 (45) | 7294 (15) | 7 | $p < 5 \times 10^{-5}$ |
| | Length of Stay (median, IQR) | 13 (5.5, 20.5) | 11 (5.0, 17.0) | 4 | $p < 5 \times 10^{-5}$ |
| | Still in hospital | 1342 (6) | 1871 (4) | 2 | $p < 5 \times 10^{-5}$ |
| | Transferred | 2052 (9) | 3457 (7) | 2 | $p < 5 \times 10^{-5}$ |
| | Discharged | 10942 (47) | 35744 (70) | 2 | $p < 5 \times 10^{-5}$ |
| | Death | 8890 (38) | 9794 (19) | 2 | $p < 5 \times 10^{-5}$ |

\*Stage 3 includes patients requiring RRT
\*\* Definitions of comorbidities, complications and outcomes from the CRFs are presented in S2 Table
RRT = Renal replacement therapy; ACEi = Angiotensin converting enzyme inhibitors; ARBs = Angiotensin II receptor blockers; NSAIDs = non-steroidal anti-inflammatories; BUN = Blood urea nitrogen; CRP = C-reactive protein; WBC = White blood cell; sCr = serum creatinine; eGFR = estimated glomerular filtration rate (estimated using the CKD-EPI equation)

**Table 3.** Characteristics of patients with AKI diagnosed using KDIGO definition versus patients diagnosed with AKI by eKDIGO only by the decrease in serum creatinine (deKDIGO).

|  |  | deKDIGO | KDIGO | Missingness (%) | P-value |
| --- | --- | --- | --- | --- | --- |
| **Total Count** |  |  |  |  |  |
|  |  | 11188 | 12704 |  |  |
| **Demographics** |  |  |  |  |  |
| | Age, yr, median (IQR) | 70 (58.5, 81.5) | 66 (56.5, 75.5) | 0 | $p < 5 \times 10^{-5}$ |
| | Female (%) | 4259 (38) | 4116 (33) | 0 | $p < 5 \times 10^{-5}$ |
| **Country Income Level, n (%)** |  |  |  |  |  |
| | High Income | 10229 (92) | 10284 (81) | 0 | $p < 5 \times 10^{-5}$ |
| | Upper Middle Income | 333 (3) | 796 (6) | 0 | $p < 5 \times 10^{-5}$ |
| | Low & low middle income | 600 (5) | 1561 (12) | 0 | $p < 5 \times 10^{-5}$ |
| **AKI Grades & RRT, n (%)** |  |  |  |  |  |
| | AKI Stage 1 | 9169 (82) | 4577 (36) | 0 | $p < 5 \times 10^{-5}$ |
| | AKI Stage 2 | 1520 (14) | 2162 (17) | 0 | $p < 5 \times 10^{-5}$ |
| | AKI Stage 3 (No RRT) | 499 (4) | 1713 (13) | 0 | $p < 5 \times 10^{-5}$ |
|  | AKI Stage 3 (with RRT) | - | 5965 (47) | 0 |  |
|  | RRT | - | 4252 (36) | 9 |  |
| **Comorbidities\*, n (%)** |  |  |  |  |  |
|  | Chronic Kidney Disease | 1797 (17) | 2262 (19) | 7 |  |
|  | Chronic Cardiac Disease | 2989 (27) | 3236 (26) | 2 |  |
| | Chronic Pulmonary Disease | 1599 (15) | 1376 (12) | 7 | $p < 5 \times 10^{-5}$ |
|  | Hypertension | 3902 (48) | 4545 (51) | 28 |  |
| | Dementia | 1316 (13) | 519 (5) | 12 | $p < 5 \times 10^{-5}$ |
| | Diabetes -type 2 | 3443 (33) | 4553 (39) | 6 | $p < 5 \times 10^{-5}$ |
|  | Liver disease | 380 (4) | 462 (4) | 4 |  |
| | Malnutrition | 284 (3) | 217 (2) | 12 | $p < 5 \times 10^{-5}$ |
| | Obesity | 1504 (16) | 2288 (22) | 18 | $p < 5 \times 10^{-5}$ |
| **Medications on Admission, n (%)** |  |  |  |  |  |
|  | NSAIDS | 627 (8) | 755 (9) | 35 |  |
|  | ACEi | 1407 (18) | 1388 (17) | 33 |  |
|  | ARBs | 926 (12) | 1016 (12) | 33 |  |
| **Signs & Symptoms on Admission, n (%)** |  |  |  |  |  |
| | Altered Consciousness / Confusion | 2510 (28) | 1785 (18) | 19 | $p < 5 \times 10^{-5}$ |
| | Diarrhea | 1885 (21) | 1855 (18) | 19 | $p < 5 \times 10^{-5}$ |
|  | Fever | 6126 (64) | 7207 (65) | 12 |  |
| | Vomiting /nausea | 1758 (20) | 1629 (16) | 19 | $p < 5 \times 10^{-5}$ |
|  | Muscle aches/joint pain | 1617 (20) | 2026 (22) | 25 |  |
|  | Headache | 743 (9) | 1036 (11) | 26 |  |
| | Sore Throat | 628 (8) | 936 (10) | 27 | $p < 5 \times 10^{-5}$ |
|  | Cough | 5979 (63) | 6982 (64) | 12 |  |
| | Shortness of breath | 6680 (70) | 8144 (73) | 12 | $p < 5 \times 10^{-5}$ |
| | Runny nose | 230 (3) | 404 (4) | 28 | $p < 5 \times 10^{-5}$ |
| **Observations on Admission, median (IQR)** |  |  |  |  |  |
|  | Temperature, °C | 37.3 (36.3, 38.3) | 37.2 (36.2, 38.2) | 3 |  |
| | Systolic BP, mmHg | 123 (107.0, 139.0) | 130 (113.5, 146.5) | 6 | $p < 5 \times 10^{-5}$ |
| | Diastolic BP, mmHg | 70 (60.5, 79.5) | 72 (62.0, 82.0) | 7 | $p < 5 \times 10^{-5}$ |
|  | Heart rate, BPM | 92 (78.5, 105.5) | 93 (79.5, 106.5) | 7 |  |
| | Respiratory rate, per min | 22 (17.5, 26.5) | 24 (19.5, 28.5) | 13 | $p < 5 \times 10^{-5}$ |
| | Oxygen saturation, % | 95 (92.0, 98.0) | 94 (91.0, 97.0) | 7 | $p < 5 \times 10^{-5}$ |
| **Laboratory results on admission, median (IQR)** |  |  |  |  |  |
| | WBC ($\times 10^9/L$) | 8.1 (5.1, 11.1) | 8.3 (5.3, 11.3) | 15 | |
| | BUN (mmol/L) | 11.6 (5.6, 17.6) | 10.1 (4.1, 16.1) | 22 | $p < 5 \times 10^{-5}$ |
|  | Potassium (mmol/L) | 4.1 (3.7, 4.5) | 4.2 (3.8, 4.6) | 15 |  |
| | CRP (mg/L) | 89 (20.0, 158.0) | 102.7 (21.2, 184.2) | 22 | $p < 5 \times 10^{-5}$ |
| | sCr, (umol/l) | 119 (80.0, 158.0) | 101 (56.5, 145.5) | 11 | $p < 5 \times 10^{-5}$ |
| | eGFR, ml/min/1.73m<sup>2</sup> | 48.6 (28.1, 69.1) | 62.1 (34.6, 89.6) | 12 | $p < 5 \times 10^{-5}$ |
| **Admission Treatment, n (%)** |  |  |  |  |  |
| | Antiviral and COVID-19 targeting agents | 2057 (22) | 3292 (31) | 16 | $p < 5 \times 10^{-5}$ |
| | Antibiotic agents | 9718 (91) | 11187 (94) | 8 | $p < 5 \times 10^{-5}$ |
| | Antifungal agents | 698 (7) | 1761 (16) | 11 | $p < 5 \times 10^{-5}$ |
| | Corticosteroids | 3248 (31) | 5305 (45) | 9 | $p < 5 \times 10^{-5}$ |
| **Complications\*, n (%)** |  |  |  |  |  |
| | Bacterial pneumonia | 1461 (15) | 2366 (22) | 16 | $p < 5 \times 10^{-5}$ |
| | Cardiac arrest | 280 (3) | 1202 (10) | 10 | $p < 5 \times 10^{-5}$ |
| | Coagulation disorder | 402 (4) | 1012 (9) | 15 | $p < 5 \times 10^{-5}$ |
| | Rhabdomyolysis | 70 (0.7) | 222 (2) | 15 | $p < 5 \times 10^{-5}$ |
| **Outcomes, n (%)** |  |  |  |  |  |
| | ICU admission | 3836 (35) | 8743 (70) | 2 | $p < 5 \times 10^{-5}$ |
| | Invasive mechanical ventilation | 2596 (24) | 7668 (63) | 7 | $p < 5 \times 10^{-5}$ |
| | Length of Stay (median, IQR) | 12 (5.5, 18.5) | 15 (6.0, 24.0) | 4 | $p < 5 \times 10^{-5}$ |
| | Still in hospital | 540 (5) | 802 (7) | 2 | $p < 5 \times 10^{-5}$ |
|  | Transferred | 924 (8) | 1128 (9) | 2 |  |
| | Discharged | 6761 (62) | 4181 (34) | 2 | $p < 5 \times 10^{-5}$ |
| | Death | 2692 (25) | 6198 (50) | 2 | $p < 5 \times 10^{-5}$ |

\* Definitions of comorbidities, complications and outcomes from the CRFs are presented in S2 Table RRT = Renal replacement therapy; ACEi = Angiotensin converting enzyme inhibitors; ARBs = Angiotensin II receptor blockers; NSAIDs = non-steroidal anti-inflammatories; BUN = Blood urea nitrogen; CRP = C-reactive protein; WBC = White blood cell; sCr = serum creatinine; eGFR = estimated glomerular filtration rate (estimated using the CKD-EPI equation)

**Table 4.** Characteristics of patients with AKI diagnosed using eKDIGO only by the decrease in serum creatinine (deKDIGO) and no AKI.

|  |  | **deKDIGO** | **No AKI** | **Missingness (%)** | **P-value** |
| --- | --- | --- | --- | --- | --- |
| **Total Count** |  |  |  |  |  |
|  |  | 11188 | 51772 |  |  |
| **Demographics** |  |  |  |  |  |
| | Age, yr, median (IQR) | 70 (58.5, 81.5) | 67 (53.5, 80.5) | 0 | $p < 5 \times 10^{-5}$ |
| | Female (%) | 4259 (38) | 22031 (43) | 0 | $p < 5 \times 10^{-5}$ |
| **Country Income Level, n (%)** |  |  |  |  |  |
| | High Income | 10229 (92) | 45686 (88) | 0 | $p < 5 \times 10^{-5}$ |
| | Upper Middle Income | 333 (3) | 4176 (8) | 0 | $p < 5 \times 10^{-5}$ |
| | Low & low middle income | 600 (5) | 1876 (4) | 0 | $p < 5 \times 10^{-5}$ |
| **AKI Grades & RRT, n (%)** |  |  |  |  |  |
|  | AKI Stage 1 | 9169 (82) | 0 (0.0) | 0 |  |
|  | AKI Stage 2 | 1520 (14) | 0 (0.0) | 0 |  |
|  | AKI Stage 3* | 499 (4) | 0 (0.0) | 0 |  |
|  | RRT | - | - | 9 |  |
| **Comorbidities\*, n (%)** |  |  |  |  |  |
| | Chronic Kidney Disease | 1797 (17) | 5433 (11) | 7 | $p < 5 \times 10^{-5}$ |
|  | Chronic Cardiac Disease | 2989 (27) | 12772 (25) | 2 |  |
|  | Chronic Pulmonary Disease | 1599 (15) | 6853 (14) | 7 |  |
| | Hypertension | 3902 (48) | 16429 (43) | 28 | $p < 5 \times 10^{-5}$ |
| | Dementia | 1316 (13) | 4315 (9) | 12 | $p < 5 \times 10^{-5}$ |
| | Diabetes -type 2 | 3443 (33) | 14051 (29) | 6 | $p < 5 \times 10^{-5}$ |
|  | Liver disease | 380 (4) | 1653 (3) | 4 |  |
| | Malnutrition | 284 (3) | 963 (2) | 12 | $p < 5 \times 10^{-5}$ |
|  | Obesity | 1504 (16) | 6404 (15) | 18 |  |
| **Medications on Admission, n (%)** |  |  |  |  |  |
|  | NSAIDS | 627 (8) | 2610 (8) | 35 |  |
| | ACEi | 1407 (18) | 5177 (15) | 33 | $p < 5 \times 10^{-5}$ |
| | ARBs | 926 (12) | 3101 (9) | 33 | $p < 5 \times 10^{-5}$ |
| **Signs & Symptoms on Admission, n (%)** |  |  |  |  |  |
| | Altered Consciousness / Confusion | 2510 (28) | 8267 (20) | 19 | $p < 5 \times 10^{-5}$ |
| | Diarrhea | 1885 (21) | 7983 (19) | 19 | $p < 5 \times 10^{-5}$ |
|  | Fever | 6126 (64) | 30320 (66) | 12 |  |
|  | Vomiting /nausea | 1758 (20) | 8099 (19) | 19 |  |
| | Muscle aches/joint pain | 1617 (20) | 8853 (23) | 25 | $p < 5 \times 10^{-5}$ |
| | Headache | 743 (9) | 5367 (14) | 26 | $p < 5 \times 10^{-5}$ |
|  | Sore Throat | 628 (8) | 3579 (9) | 27 |  |
|  | Cough | 5979 (63) | 29549 (65) | 12 |  |
| | Runny nose | 230 (3) | 1547 (4) | 28 | $p < 5 \times 10^{-5}$ |
| **Observations on Admission, median (IQR)** |  |  |  |  |  |
|  | Temperature, °C | 37.3 (36.3, 38.3) | 37.3 (36.8, 37.8) | 3 |  |
| | Systolic BP, mmHg | 123 (107.0, 139.0) | 130 (115.0, 145.0) | 6 | $p < 5 \times 10^{-5}$ |
| | Diastolic BP, mmHg | 70 (60.5, 79.5) | 75 (66.0, 84.0) | 7 | $p < 5 \times 10^{-5}$ |
| | Heart rate, BPM | 92 (78.5, 105.5) | 90 (77.5, 102.5) | 7 | $p < 5 \times 10^{-5}$ |
| | Respiratory rate, per min | 22 (17.5, 26.5) | 21 (17.0, 25.0) | 13 | $p < 5 \times 10^{-5}$ |
| | Oxygen saturation, % | 95 (92.0, 98.0) | 95 (92.5, 97.5) | 7 | $p < 5 \times 10^{-5}$ |
| **Laboratory results on admission, median (IQR)** |  |  |  |  |  |
| | WBC ($\times 10^9/L$) | 8.1 (5.1, 11.1) | 7 (4.5, 9.5) | 15 | $p < 5 \times 10^{-5}$ |
| | BUN (mmol/L) | 11.6 (5.6, 17.6) | 6.4 (3.9, 8.9) | 22 | $p < 5 \times 10^{-5}$ |
| | Potassium (mmol/L) | 4.1 (3.7, 4.5) | 4.1 (3.7, 4.4) | 15 | $p < 5 \times 10^{-5}$ |
| | CRP (mg/L) | 89 (20.0, 158.0) | 69 (12.5, 125.5) | 22 | $p < 5 \times 10^{-5}$ |
| | sCr (umol/L) | 119 (80.0, 158.0) | 80 (63.0, 97.0) | 11 | $p < 5 \times 10^{-5}$ |
| | eGFR, (ml/min/1.73m<sup>2</sup>) | 48.6 (28.1, 69.1) | 80.3 (61.8, 98.8) | 12 | $p < 5 \times 10^{-5}$ |
| **Admission Treatment, n (%)** |  |  |  |  |  |
|  | Antiviral and COVID-19 targeting agents | 2057 (22) | 9145 (21) | 16 |  |
| | Antibiotic agents | 9718 (91) | 40430 (86) | 8 | $p < 5 \times 10^{-5}$ |
| | Antifungal agents | 698 (7) | 2564 (6) | 11 | $p < 5 \times 10^{-5}$ |
| | Corticosteroids | 3248 (31) | 11905 (25) | 9 | $p < 5 \times 10^{-5}$ |
| **Complications\*\*, n (%)** |  |  |  |  |  |
| | Bacterial pneumonia | 1461 (15) | 5761 (13) | 16 | $p < 5 \times 10^{-5}$ |
|  | Cardiac arrest | 280 (3) | 988 (2) | 10 |  |
| | Coagulation disorder | 402 (4) | 1321 (3) | 15 | $p < 5 \times 10^{-5}$ |
|  | Rhabdomyolysis | 70 (0.7) | 177 (0.4) | 15 |  |
| **Outcomes, n (%)** |  |  |  |  |  |
| | ICU admission | 3836 (35) | 11652 (23) | 2 | $p < 5 \times 10^{-5}$ |
| | Invasive mechanical ventilation | 2596 (24) | 7294 (15) | 7 | $p < 5 \times 10^{-5}$ |
| | Length of Stay (median, IQR) | 12 (5.5, 18.5) | 11 (5.0, 17.0) | 4 | $p < 5 \times 10^{-5}$ |
| | Still in hospital | 540 (5) | 1871 (4) | 2 | $p < 5 \times 10^{-5}$ |
| | Transferred | 924 (8) | 3457 (7) | 2 | $p < 5 \times 10^{-5}$ |
| | Discharged | 6761 (62) | 35744 (70) | 2 | $p < 5 \times 10^{-5}$ |
| | Death | 2692 (25) | 9794 (19) | 2 | $p < 5 \times 10^{-5}$ |

\*Does not include RRT
\*\* Definitions of comorbidities, complications and outcomes from the CRFs are presented in S2 Table
RRT = Renal replacement therapy; ACEi = Angiotensin converting enzyme inhibitors; ARBs = Angiotensin II receptor blockers; NSAIDs = non-steroidal anti-inflammatories; BUN = Blood urea nitrogen; CRP = C-reactive protein; WBC = White blood cell; sCr = serum creatinine; eGFR = estimated glomerular filtration rate (estimated using the CKD-EPI equation)

A logistic regression was fitted to assess the association between eKDIGO AKI with in-hospital mortality. A t-test with a significance threshold of 0.001 was used to assess the significance of predictors. MICE imputation was used to address variable missingness and a sensitivity analysis showing the results without imputation can be found in S4 Table. Adjustments were made for factors indicating disease severity such as: admission to ICU; need for mechanical ventilation; corticosteroid and antifungal treatment; complicating factors such as bacterial pneumonia, cardiac arrest, coagulation disorders and rhabdomyolysis. Adjustment was also made for factors known to increase the susceptibility to AKI such as age, sex, diabetes, chronic cardiac disease, chronic pulmonary disease, chronic kidney disease (CKD), hypertension, obesity, and use of renin–angiotensin system blockers (RAS blockers) before admission [5].

The relationship between eKDIGO AKI and in-hospital death and discharge was described with a survival curve approximated using the Aalen–Johansen estimator, a multistate version of the Kaplan-Meier estimator [14]. The follow-up period began on the day of hospital admission and ended on the day of either discharge or death, or 28 days post admission if no event had occurred. Discharge from hospital was considered an absorbing state (once discharged there was no readmission or death).

All statistical analyses were performed using the R statistical programming language, version 4.0.2 [15, 16]. This study is reported as per the Strengthening the Reporting of Observational Studies in Epidemiology (STROBE) guideline (S5 Table).

## Results

From February 15^th^, 2020, to February 1^st^, 2021, data were collected for 418,111 individuals admitted to hospital with clinically suspected or laboratory confirmed SARS-COV-2 infection from 1,609 sites and 54 countries. Of these, 75,670 were used as the analysis cohort (Fig. 1). The median length of admission was 12 days (IQR 7, 20 days). Missing data was less than 10% for most variables—although averaging 20% for symptoms on admission—and distributed evenly between groups for those with higher missingness levels (S3 Table).

### Incidence, staging and timing of peak AKI

With the KDIGO definition 12,704 (16.8 %) patients were identified as having AKI during their admission. Using the extended KDIGO definition a total of 23,892 (31.6%) patients were diagnosed with AKI. A breakdown of the top ten contributing countries for patients in each group can be found in S1 Fig. The peak stages of AKI with KDIGO and eKDIGO respectively were: stage 1 - 36% and 58%, stage 2 - 17% and 15% and stage 3 - 47% and 27% with a total of 4,252 patients (overall 5.6 % of all patients) requiring acute dialysis (Fig 3). Peak sCr occurred more frequently on days 3 and 6 from admission and diminished significantly after day 10 using a KDIGO definition. With the extended definition an additional 4,019 patients had AKI on day 3 (70% of all AKI diagnosed on that day) and 1,808 on day 6 (64% of all AKI on that day) (Fig 4).

**Fig 3.**
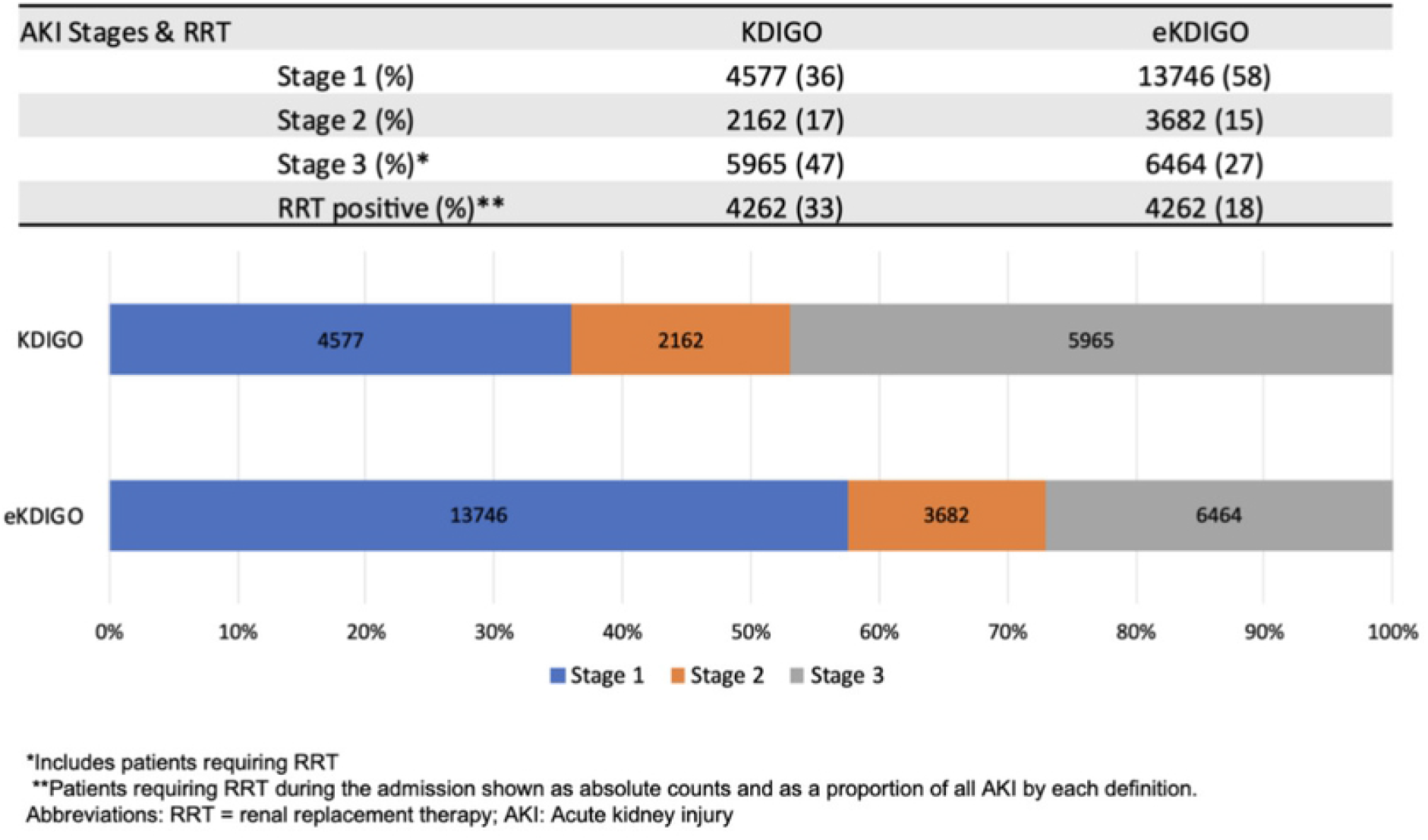
Staging of AKI using KDIGO and eKDIGO definitions.

**Fig 4.**
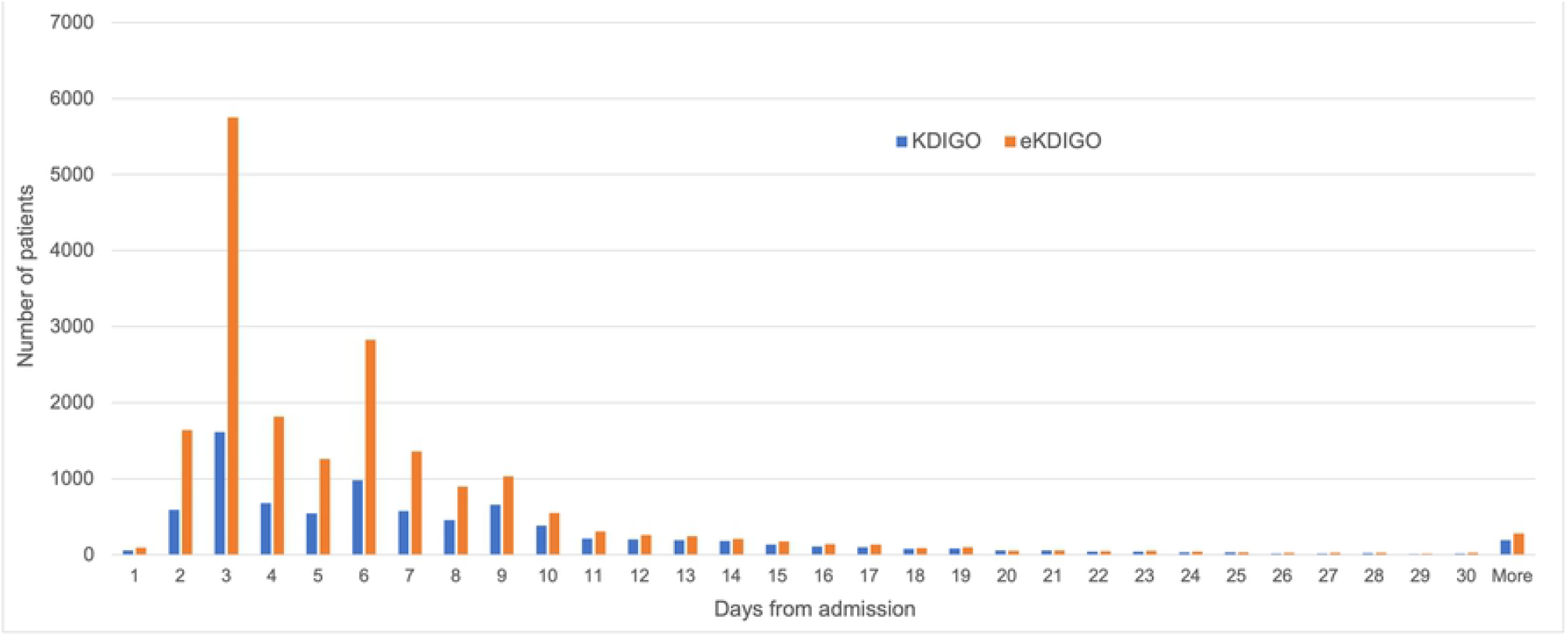
Day of peak AKI using KDIGO and eKDIGO definitions.

### Demographic and clinical characteristics

Baseline characteristics at hospital admission, acute interventions, complications and outcomes for patients with AKI diagnosed by eKDIGO vs no AKI, KDIGO vs deKDIGO and deKDIGO vs no AKI are provided in Tables 2, 3 and 4 respectively. A majority of patients from all groups were from high income countries with the highest proportion from LLMICs in the KDIGO group (12% vs 5% in eKDIGO and 9% deKDIGO). Significantly more stage 1 AKI could be seen in the deKDIGO than in KDIGO patients (82% vs 36%) while more severe forms of AKI (stage 3), were dominant in the KDIGO group even after excluding RRT patients (13% vs 4%).

Chronic kidney disease (CKD), hypertension, type 2 diabetes and obesity were significantly more common in patients who developed AKI. The use of ACE inhibitors and angiotensin receptor blockers (ARB) medications were more common in the KDIGO group as compared to both the deKDIGO group and the no AKI group. Similarly, administration of antifungal agents and corticosteroids were more common among KDIGO-diagnosed than deKDIGO AKI patients and patients without AKI. Signs, symptoms and observations at admission were very similar in all groups. At presentation, patients with eKDIGO AKI had a higher blood urea nitrogen (median 10.9 vs 6.4 mmol/l), C-reactive protein (median 95.3 vs 69 mg/L) and creatinine (110 umol/l vs 80 umol/l) and lower estimated glomerular filtration rate (eGFR, estimated with CKD-EPI equation) (54 ml/min/1.73m^2^ vs 80 ml/min/1.73m^2^) compared to those without AKI. Renal function on admission was worse in the deKDIGO group than in KDIGO patients (eGFR 48 ml/min vs 62 ml/min). In general, patients with AKI were more likely to have complications during their hospital stay.

### Clinical outcomes

Patients who developed AKI using eKDIGO were more likely to be admitted to the ICU (54%), require invasive mechanical ventilation (45%) and die during their admission (38%) compared to patients without AKI. After adjusting for disease severity, this group of patients had a higher risk of in-hospital death (OR: 1.77, 95% CI: 1.7-1.85, p-value < 0.001) (Table 5) which is further illustrated in the survival curves shown in Fig 5. Patients in the deKDIGO group appeared to have better outcomes and less mortality than those diagnosed by KDIGO criteria, but still had significantly worse outcomes and mortality than patients with no AKI (Tables 3 and 4).

**Table 5.** Logistic regression fitted to assess the association between eKDIGO AKI with in-hospital mortality.

| Variable | Odds Ratio | 95% Confidence Interval |  | P value |
| --- | --- | --- | --- | --- |
|  |  | Lower | Upper |  |
| (Intercept) | 0.051 | 0.048 | 0.054 | < 0.001 |
| AKI eKDIGO | 1.776 | 1.705 | 1.85 | < 0.001 |
| Female | 0.78 | 0.749 | 0.812 | < 0.001 |
| Age 18-65 (ref) | 1.0 | - | - | - |
| Age 65 - 85 | 3.969 | 3.778 | 4.17 | < 0.001 |
| Age 85+ | 7.773 | 7.285 | 8.294 | < 0.001 |
| Chronic Kidney Disease | 1.317 | 1.248 | 1.39 | < 0.001 |
| Chronic Cardiac Disease | 1.331 | 1.271 | 1.393 | < 0.001 |
| Chronic Pulmonary Disease | 1.514 | 1.436 | 1.595 | < 0.001 |
| Hypertension | 0.955 | 0.896 | 1.019 | 0.153 |
| Obesity | 0.904 | 0.841 | 0.972 | 0.008 |
| Diabetes Type 2 | 1.153 | 1.096 | 1.214 | < 0.001 |
| Pre-admission ACEi & ARBs | 0.867 | 0.812 | 0.926 | < 0.001 |
| Treatment with corticosteroids | 1.14 | 1.091 | 1.192 | < 0.001 |
| Treatment with antifungal agents | 1.218 | 1.138 | 1.305 | < 0.001 |
| ICU Admission | 1.585 | 1.482 | 1.695 | < 0.001 |
| Mechanical Ventilation | 2.188 | 2.037 | 2.349 | < 0.001 |
| Cardiac Arrest | 19.261 | 16.821 | 22.055 | < 0.001 |
| Bacterial Pneumonia | 1.222 | 1.16 | 1.287 | < 0.001 |
| Coagulation disorder | 1.372 | 1.253 | 1.501 | < 0.001 |
| Rhabdomyolysis | 1.17 | 0.947 | 1.444 | 0.145 |

MiCE imputation used for variable missingness
ACEi: Angiotensin converting enzyme inhibitors; ARBs: Angiotensin receptor blockers

**Fig 5.**
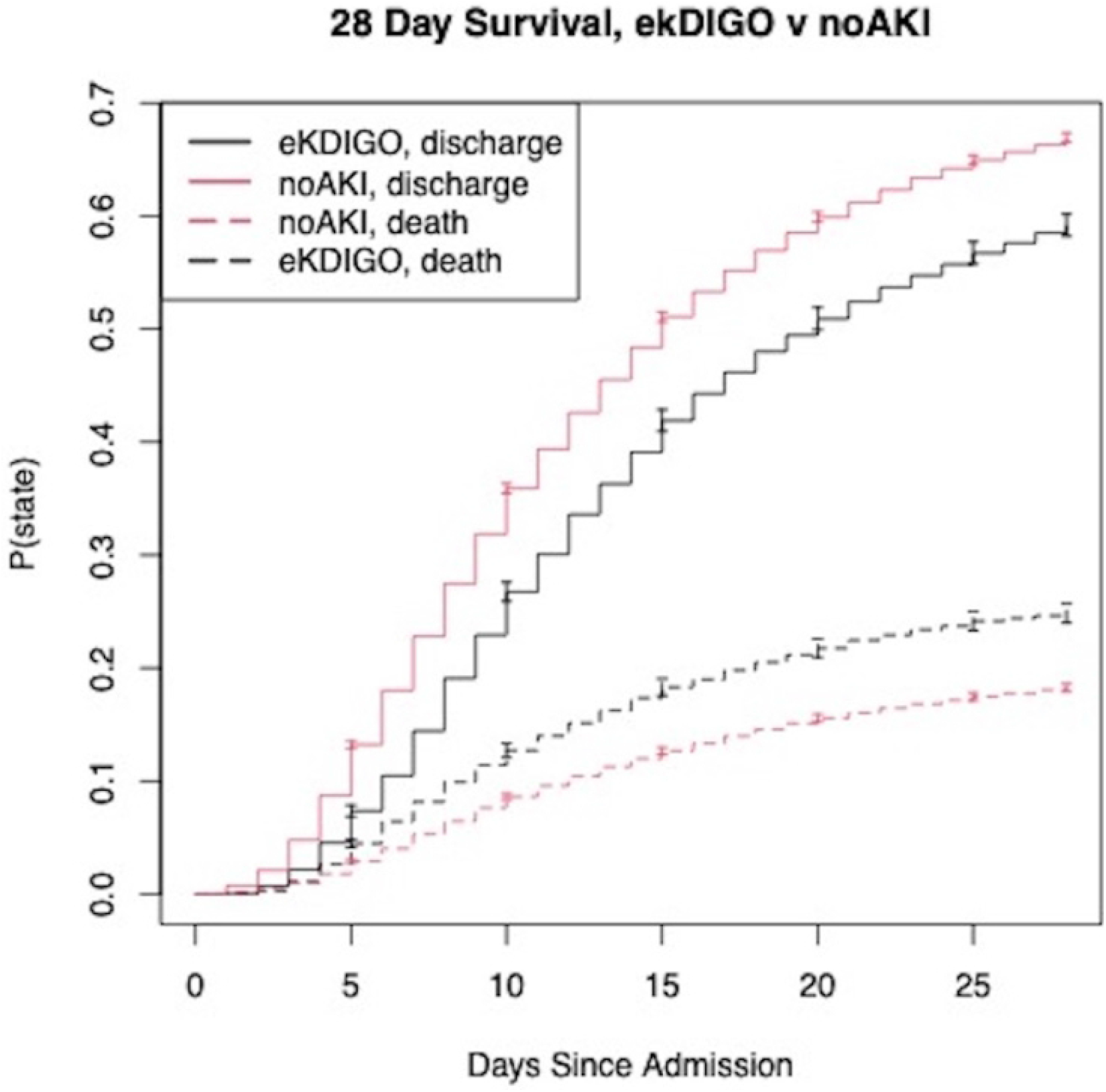
Aalen-Johansen survival plot. Outcomes among patients with AKI diagnosed using eKDIGO criteria and no AKI. Confidence bars are used to illustrate a 95% confidence interval.

## Discussion

In the largest, multinational cohort of hospitalized patients with COVID-19 it was found that an extended KDIGO criteria for the diagnosis of AKI, which includes a fall in sCr during admission, identified almost twice as many cases of AKI than the traditional KDIGO definition. The majority of these additional cases were stage 1 AKI, occurring early in the admission, supporting the hypothesis that they may represent recovering CA-AKI. This group had comparatively worse outcomes than patients without AKI, making their identification and exploration in future studies enormously important.

The estimated incidence of KDIGO AKI, 16.8%, is consistent with that reported in the first systematic review of AKI in COVID-19 patients [17], while the incidence of eKDIGO AKI fits those studies from the larger NYC cohorts which had similar rates of ICU admission [2, 18]. The mortality rate of 50% among KDIGO-diagnosed AKI patients falls within the range (34 to 50%) reported in previous studies using the same AKI definition [1, 2, 18-20]. While the inability to exclude readmitted patients may have introduced a degree of survival bias, the fact that readmission rates of less than 3% are seen in other large studies, suggests that the effect of this bias is likely to be relatively small [2, 18, 20].

Unsurprisingly, patients identified in the present study as having AKI—by either definition—were more likely to have chronic kidney disease, hypertension, and type 2 diabetes mellitus, be on an ACEi or ARB and generally have more medical complications during their admission than patients who did not develop AKI.

Interestingly, the admission eGFR, serum creatinine, and blood urea levels of the eKDIGO AKI population, and specifically those in the deKDIGO group, demonstrated significant impairment early in the admission, suggestive of community acquired AKI which would otherwise have gone unrecognized. While these patients had comparatively milder AKI and disease severity than patients in the KDIGO group, they nonetheless incurred significantly more morbidity and mortality than patients without AKI, even after adjusting for confounding factors. With regards to the increased prevalence of stage 1 AKI using the eKDIGO definition, there is growing evidence that even mild episodes of AKI may contribute to the development of chronic kidney disease [21-23]. This raises the important question of whether this new group of COVID-19 AKI patients would benefit from early management strategies to improve long term outcomes. Such measures are typically simple—management of fluid balance and removal of nephrotoxic medication for example—and readily implementable, even in resource-poor environments. A follow-up study, similar to the 0by25 feasibility study [7], may be warranted to explore such questions.

The earlier timing of peak AKI in the hospital stay and large proportion of stage 1 cases in the eKDIGO group suggests several possible etiologies. It may point to a pre-renal pattern of injury occurring in the setting of dehydration from gastrointestinal fluid losses, fever and anorexia—a finding supported by the identification of acute tubular injury in autopsy studies of patients with COVID-19 [24]. However, it is also possible that a proportion of these additional, milder, cases of AKI, captured by down trending sCr, are a consequence of early rehydration of patients with either previously normal kidney function or CKD. While the reduced admission eGFR of this group (median 54 ml/min/1.73 m^2^) makes the former less likely, pre-admission sCr measurements would be required to reliably identify the latter. It is reassuring that the proportion of reported CKD in the KDIGO and eKDIGO groups is very similar (19% and 18% respectively).

It is interesting to consider to what extent the large number of additional cases captured by the extended KDIGO definition are a COVID-specific consequence. While meta-analysis suggests global estimates of AKI incidence in adult hospitalized patients range between 3 and 18% [25], there are no current estimates of global AKI incidence according to the eKDIGO definition. Moving forward, evaluation of the eKDIGO definition for the diagnosis of AKI in various hospital and community settings will be needed to shed light on whether our findings are particular to a COVID-affected population. In this context, it should be noted that approximately 20% of the analysis cohort had a diagnosis of COVID-19 made on clinical grounds, most likely due to testing shortages and high resource demands during surge phases of the pandemic. While this may have resulted in the potential inclusion of patients with other respiratory illnesses, given that other common respiratory illnesses were notably less prevalent during the pandemic [26, 27], it is plausible that a significant proportion of these clinically diagnosed patients did in fact have COVID-19.

This study has some key limitations. The exclusion of patients without two sCr measurements may have introduced a degree of selection bias. This could be responsible for the absence of expected geographical differences found between the eKDIGO and no AKI groups and may also have resulted in an underestimation of AKI cases by both definitions [6]. The lack of a time-standardized collection of sCr across all sites also represents a limitation of the study. Patients having more frequent sCr collections may represent a population with more severe illness in whom AKI would be more readily detected, therefore affecting the overall AKI incidence rates and potentially generating a negative survival bias. Nevertheless, it is reassuring that the number of AKI cases are a small proportion of the total sCr collected on any given day (<18%) (S2 Fig), suggesting that the bias introduced by ad hoc sampling was low. The lack of standardization in sCr collection may have also affected the reporting of time to peak AKI, the magnitude of peak AKI reached in each individual patient and, in those experiencing both a rise and fall in sCr during their admission, whether AKI was captured during the former phase (KDIGO) or the latter (eKDIGO).

With regards to the distinction between community and hospital acquired AKI, often a 48 hr threshold is used to identify CA-AKI [28]. Such a definition would preclude many patients in this study who were identified as having AKI on day 3 of admission. It is worth noting that these patients would be identified as CA-AKI (or transient hospital-associated AKI) by the definition proposed by Warnock et al which integrates serum creatinine trajectories and doesn’t adhere to the somewhat arbitrary 48hr cutoff [29]. Whether or not the additional cases of AKI captured by eKDIGO are truly reflective of CA-AKI will ultimately require studies that assess this population in a variety of community settings.

To our knowledge, this is the first study to systematically examine an extended KDIGO definition for the identification of AKI against the traditional KDIGO criteria in hospitalized COVID-19 patients. Our population is, as far as we know, the largest and only multinational cohort of patients with COVID-19 from all income country levels. The use of an extended KDIGO definition to diagnose AKI in this population resulted in a significantly higher incidence rate compared to traditional KDIGO criteria. These additional cases of AKI appear to be occurring in the community or early in the hospital admission and are associated with significantly worse outcomes, highlighting the importance of examining their role and long-term impact in future studies.

## Data Availability

The data that underpin this analysis are highly detailed clinical data on individuals hospitalised with COVID-19. Due to the sensitive nature of these data and the associated privacy concerns, they are available via a governed data access mechanism following review of a data access committee. Data can be requested via the IDDO COVID-19 Data Sharing Platform (www.iddo.org/covid-19). The Data Access Application, Terms of Access and details of the Data Access Committee are available on the website. Briefly, the requirements for access are a request from a qualified researcher working with a legal entity who have a health and/or research remit a scientifically valid reason for data access which adheres to appropriate ethical principles. The full terms are at https://www.iddo.org/document/covid-19-data-access-guidelines. A small subset of sites who contributed data to this analysis have not agreed to pooled data sharing as above. In the case of requiring access to these data, please contact the ISARIC team at in the first instance who will look to facilitate access.

https://isaric.net/ccp/

## Supplementary material

**S1 Statement**. Study ethics approval

**S1 Table**. Definitions used for clinical COVID-19

**S2 Table**. Definition of comorbidities, complications and outcomes from the ISARIC case report forms (CRF)

**S3 Table**. Distribution of missingness information between eKDIGO and No AKI patients.

**S4 Table**. Logistic regression fitted to assess the association between eKDIGO AKI with in-hospital mortality without MICE imputation for variable missingness

**S1 Fig**. Breakdown of top contributing countries for patients diagnosed with AKI by KDIGO definition (A) and from deKDIGO group (B)

**S2 Fig**. Number of AKI cases by AKI definition (A = KDIGO and B = eKDIGO) as a proportion of total number of serum creatinines collected each day

**S5 Table**. STROBE (Strengthening the reporting of observational studies in epidemiology) checklist

